# Analysis and evaluation of blood screening testing strategies in Nagqu, Tibet: one of the highest cities in world

**DOI:** 10.1101/2023.11.08.23297532

**Authors:** Liang Zang, Lei Zhou, Yaxin Fan, Xiaohua Liang, Ji Duo, Hao Lin, Rui Bai, Mei Yang, Chao Dan

**Affiliations:** Dalian Blood Center, Dalian, Liaoning, China; Nagqu Central Blood Station, Nagqu, Tibet, China

## Abstract

The aim of this study was to evaluate the performance of blood screening strategies in Nagqu City, with a focus on serological and nucleic acid testing effectiveness. Analysis of blood donation data from 2017 to 2023 revealed a gradual increase in local blood donation rates, but the rate of unqualified donations remained higher than the national average. Serological performance verification showed good repeatability and precision, but improvements are needed in the accuracy of certain markers. Nucleic acid performance verification demonstrated excellent detection capability for low viral load HBV infections. The conclusion highlights significant progress in the screening strategies of Nagqu City, with the nucleic acid testing system showing outstanding performance. The findings provide valuable insights for blood screening in other high-altitude regions.

## Introduction

Nagqu City, nestled in the northern part of the Tibet Autonomous Region in China on the awe-inspiring Tibetan Plateau, stands as one of the highest cities on Earth, boasting an average elevation of approximately 4,500 meters (14,764 feet) above sea level[1]. Its population of 504,838 has transformed the city into a crucial trade and commercial hub, fostering vital connections between various Tibetan regions and neighboring countries[2]. Nevertheless, the delivery of healthcare services, especially blood donation screening, faces formidable challenges due to the city’s geographical remoteness, limited infrastructure, and the demanding high-altitude conditions it presents[3].

A significant stride towards improving blood donation and healthcare services in the region occurred in 2017 with the establishment of a blood station in Nagqu City. However, this unique city’s population demographics and high-altitude environment introduce distinct challenges that demand careful consideration for ensuring effective and reliable blood screening and donation procedures[4–9].

Given Nagqu City’s remote location and limited infrastructure, the validation and evaluation of laboratory blood testing systems, including serological testing and nucleic acid testing (NAT), assume paramount importance[10]. This validation entails assessing the accuracy, sensitivity, and specificity of screening methods for detecting infectious diseases and Transfusion-Transmissible Infections (TTIs), such as Hepatitis B virus (HBV), Hepatitis C virus (HCV), Human Immunodeficiency Virus (HIV), and Syphilis. Strict adherence to relevant regulations, such as the ‘China Blood Bank Quality Management Standards,’ ‘China Blood Bank Laboratory Quality Management Standards,’ and ‘China Blood Technical Operating Procedures (2019 edition)’ [11–13], ensures the provision of a safe and adequate blood supply to meet the healthcare needs of the local population.

In pursuit of evaluating the efficacy of current blood screening strategies, this study meticulously analyzes blood screening results obtained from the Nagqu City blood station laboratory over several years, with a primary focus on comprehending the epidemiological trends of TTIs among the local donor population. The profound insights into the prevalence and distribution of TTIs will facilitate the identification of potential risks and areas for refining screening strategies, thereby ensuring a safe blood supply and mitigating the transmission of infectious diseases[14]. Furthermore, the study scrutinizes the External Quality Assessment (EQA) results of the Nagqu blood station to assess the laboratory’s performance and accuracy in conducting blood screening tests. This rigorous evaluation bolsters the reliability and consistency of screening procedures, while also pinpointing any factors that could impact result quality.

Through this comprehensive evaluation of current blood screening strategies, the ultimate aim of this study is to refine and optimize the screening protocols, ensuring their unwavering effectiveness and reliability. In doing so, this endeavor seeks to enhance the safety of the blood supply in Nagqu City and potentially engender valuable insights applicable to other high-altitude regions worldwide grappling with similar challenges.

## Materials and methods

### Sample Collection

A total of 3,585 blood samples were collected from donors in Nagqu City using standard procedures from September 1, 2017 to July 30, 2023. The informed consents were duly obtained from all participants. Furthermore, ethical approval was obtained from the Ethics Committee of the Dalian Blood Center (LL-202202). The screening process involved specific testing equipment for serological and nucleic acid testing.

The serological test performance verification (PV) serum panels included HBsAg Serum Panel, HCV Ab Serum Panel, HIV-1 Ab Serum Panel, and TP Ab Serum Panel. Each panel consisted of 20 positive samples, 20 negative samples, 5 analytical sensitivity samples, and a set of precision verification samples. The HIV-1 P24 Serum Panel included 10 positive samples, 20 negative samples, 10 analytical sensitivity samples, and a set of precision verification samples. All serum panels were provided by Control Standard Co., Ltd.(Beijing, China). Blood Screening Four-Item Quality Control Samples and Negative Quality Control Samples were provided by Control Standard Co., Ltd.(Beijing, China).

The NAT PV serum panels included the NAT Serum Panels C to G (analytical sensitivity verification panels) included a total of 30 samples, with HBV genotype C samples (C2 to C6) having concentrations of 25, 12, 6, 3, and 1.5 IU/mL, HCV subtype 2a samples (D2 to D6) having concentrations of 170, 85, 40, 20, and 10 IU/mL, HIV genotype CRF07_BC samples (E2 to E6) having concentrations of 210, 105, 50, 25, and 12.5 IU/mL, HCV subtype 1b samples (F2 to F6) having concentrations of 140, 70, 35, 17, and 8.5 IU/mL, and HCV subtype 6a samples (G2 to G6) having concentrations of 220, 110, 55, 27, and 13 IU/mL. The panel also included 5 negative control samples. The NAT evaluation serum panel A consisted of a total of 46 samples, with each numbered as 3 vials, and each sample having 3 mL. All NAT serum panels were provided by the National Center for Clinical Laboratories (NCCL).

### Testing Equipment and Reagents

For serological testing, the ELISA method was used with the Aikang URANUS AE 158 enzyme immunoassay system produced by Shenzhen Aikang Biotechnology Co., Ltd. For nucleic acid testing, the DaAn Gene DA3500S Nucleic Acid Extraction System from Guangzhou, China, and the Aoyu AGS4800 PCR Amplification System from Hangzhou, China, were employed.

Various diagnostic kits were used for specific tests:

HBsAg (Hepatitis B Surface Antigen) diagnostic kit: InTec Products, Inc. (Xiamen, China), WANTAI BioPharm, Co., Ltd. (Beijing, China)

HCV (Hepatitis C Virus) antibody diagnostic kit: WANTAI BioPharm, Co., Ltd. (Beijing, China), Livzon Pharmaceutical Group Inc. (Zhuhai, China)

HIV (Human Immunodeficiency Virus) antibody diagnostic kit: WANTAI BioPharm, Co., Ltd. (Beijing, China), Livzon Pharmaceutical Group Inc. (Zhuhai, China)

Treponema pallidum (Syphilis) antibody diagnostic kit: InTec Products, Inc. (Xiamen, China), WANTAI BioPharm, Co., Ltd. (Beijing, China)

Nucleic Acid Extraction: DaAn Blood Screening HBV, HCV, and HIV-1 Virus Nucleic Acid Test Kit from DAAN Gene (Guangzhou, China)

All testing reagents and kits were selected based on their suitability for detecting specific markers and pathogens related to Hepatitis B, Hepatitis C,

HIV, and syphilis. The testing procedures and methods followed the instructions provided by the manufacturers.

### Screening Strategy

The serological markers were tested using different reagents, and each marker was tested once. Single-Side Reactivity: If a single-side reagent reaction was observed, a necessary repeat test was conducted to confirm the result. If the repeated test still showed reactivity, the sample was considered unqualified for single-side reactivity. Dual-Side Reactivity: If both side reagents showed reactivity simultaneously, the sample was considered unqualified for both sides. A non-reactive result in the nucleic acid test was considered qualified, while a reactive result was deemed unqualified.

Both serological testing and NAT were conducted simultaneously, and blood donations with qualified results for both tests were released.

### Serological Test Validation Procedure

#### Reproducibility

Each Blood Screening Four-Item Quality Control Sample and Blood Screening Negative Quality Control Sample were tested 10 times. All 20 test results can not appear more than 1 negative/positive control does not meet the manufacturer’s requirements.

#### Precision

The precision verification samples of PV serum panels was tested within one day. The CV of variation of intra-batch imprecision should be less than 15%.

#### Sensitivity and Specificity

The PV serum panels were applied and tested according to the instructions. All samples within the blood screening panel were tested within one day. The sensitivity and specificity are calculated based on the detection results.

#### Compliance

The test results of laboratory samples participating in NCCL were analyzed, the consistency of laboratory test results and NCCL target value results was compared, and the coincidence rate of detection method was calculated.

#### Determination of Limit of Detection (LOD)

Sensitivity samples from PV serum panels were subjected to serial dilution. For each individual assay, three different concentrations were prepared and subjected to 20 repeated tests. The LOD for each assay was calculated as the lowest concentration at which 95% of the results yielded a Signal-to-Cut-off ratio (S/CO) ≥1.

### NAT Validation Procedure

#### Analytical Sensitivity

The laboratory evaluated the analytical sensitivity using NAT Serum Panels C to G provided by NCCL. The samples covered the minimum detection limits declared in the reagent instructions (HBV/HCV/HIV ≤ 100 IU/mL). Before testing, the samples were thawed at room temperature and mixed thoroughly by vortexing for 30 seconds or inverting 30 times. Each sensitivity verification panel was tested over three days, with each concentration tested in at least 6 replicates per day, totaling no less than 20 replicates over the three days. SPSS 20.0 was used for PROBIT analysis to assess the LOD and the two-sided 95% CI for each virus.

#### Precision

The laboratory assessed precision using standard materials with concentrations less than 3 times the LOD for HBV DNA(25 IU/mL), HCV RNA (170 IU/mL), HIV-1 RNA (105 IU/mL) and negative controls provided by the NCCL. Each material was tested 20 times, and the results were collected to calculate the system’s precision.

#### Cross-Contamination

To evaluate cross-contamination, five positive standard materials (C2 to G2) from the Analytical Sensitivity Serum Panel were placed together with five negative standard materials from the NAT Serum Panels in a single-test mode using the sample rack. The laboratory analyzed the detection results of both positive and negative standard materials to assess any cross-contamination.

#### Consistency

The laboratory analyzed the test results of laboratory samples participating in the NCCL EQA. The coincidence rate between the laboratory’s test results and the NCCL results was calculated to evaluate the system’s consistency.

#### Detecting low viral load HBV infections

46 samples with low concentration of HBV DNA from NAT Serum A provided by the NCCL. The samples were remelted at room temperature before detection, swirled for 30 seconds or reversed for 30 times, thoroughly mixed, and centrifuged for 1000-1500g for 5 minutes. All samples were tested by single-sample test, the first test was only once, and the second test was double-hole retest.

## Results

### Innovations and Challenges in Blood Donation

Despite facing geographical remoteness and limited infrastructure, Nagqu City has made remarkable efforts to overcome challenges in healthcare delivery and blood donation. Innovations in healthcare include the implementation of telemedicine, providing remote consultations and medical services, along with the deployment of mobile healthcare units to extend essential care to remote communities. Community education and engagement have played a pivotal role in promoting blood donation, with awareness campaigns and blood drives leading to a significant increase in donors. As indicated in Table 1, these initiatives have contributed to an overall rise in blood donations over the years, showcasing the active participation of the local population in blood donation. However, despite progress in healthcare and blood donation, challenges persist. Factors such as religious beliefs among ethnic minorities and seasonal weather conditions contribute to blood shortages in Nagqu City. Severe weather conditions from October to April, including snowstorms, windstorms, hailstorms, and flooding, significantly impact the accessibility and availability of blood donations. These unique challenges necessitate further efforts to address the issue of blood shortages while considering the specific religious and climatic factors in the region.

**Table 1.** Number of Donations and Number (Rate) unqualified Donations in Nagqu City from 2017 to 2023.

| Year | Donations | Serological Test |  |  |  |  | NAT |  |  |
| --- | --- | --- | --- | --- | --- | --- | --- | --- | --- |
|  |  | ALT | HBsAg | Anti-HCV | Anti-HIV | Anti-TP | HBV-DNA | HCV-RNA | HIV-RNA |
| 2017 | 35 | 3 (8.57%) | 1 (2.86%) | 1 (2.86%) | 1 (2.86%) | 0 | 0 | 0 | 0 |
| 2018 | 186 | 11 (5.91%) | 0 | 0 | 2 (1.08%) | 2 (1.08%) | 3 (1.61%) | 0 | 0 |
| 2019 | 288 | 17 (5.90%) | 5 (1.74%) | 0 | 1 (0.35%) | 1 (0.35%) | 3 (1.04%) | 0 | 0 |
| 2020 | 764 | 36 (4.71%) | 4 (0.52%) | 0 | 0 | 13 (1.70%) | 4 (0.52%) | 0 | 0 |
| 2021 | 1117 | 48 (4.30%) | 5 (0.45%) | 1 (0.09%) | 2 (0.18%) | 8 (0.72%) | 6 (0.54%) | 0 | 0 |
| 2022 | 695 | 15 (2.16%) | 6 (0.86%) | 0 | 1 (0.14%) | 7 (1.01%) | 7 (1.01%) | 1 (0.14%) | 0 |
| 2023 | 500 | 13 (2.60%) | 3 (0.60%) | 0 | 1 (0.20%) | 8 (1.60%) | 5 (1.00%) | 0 | 1 (0.20%) |
| Total | 3585 | 143 (3.99%) | 24 (0.67%) | 2 (0.06%) | 8 (0.22%) | 39 (1.09%) | 28 (0.78%) | 1 (0.03%) | 1 (0.03%) |
| National Average Rate<br>(2021) |  | 0.75% | 0.46% | 0.13% | 0.12% | 0.30% |  | 0.13% |  |

Note: The data access period was from August 1, 2023. The data was collected within the timeframe from September 1, 2017, to June 31, 2023. After data collection, authors had access to information that could identify individual participants.

Table 1 and Fig 1 provide data on the number and rate of unqualified donations over the years, demonstrating the effectiveness of serological testing strategies. The rate of unqualified donations has generally decreased over time, indicating an improvement in the effectiveness of serological testing strategies. However, when compared to the national average rates from the 2021 National Research on Blood Screening, Nagqu City’s rates of unqualified donations for various blood screening markers consistently exceed the national average. This highlights the challenge of ensuring a safe and adequate blood supply in Nagqu City, given the high prevalence of TTIs, particularly HBV.

**Fig 1.** Time trend for prevalence of ALT, HBV, HCV, HIV, and syphilis among donations in Nagqu of China, 2017–2023

### Performance Verification of Blood Screening

To further analyze the performance of blood screening strategies in Nagqu City, we conducted performance verification for both serological and nucleic acid testing procedures. The results, presented in Tables 2, 3 and 4, provide valuable insights into the reliability and accuracy of the testing methods.

**Table 2.**
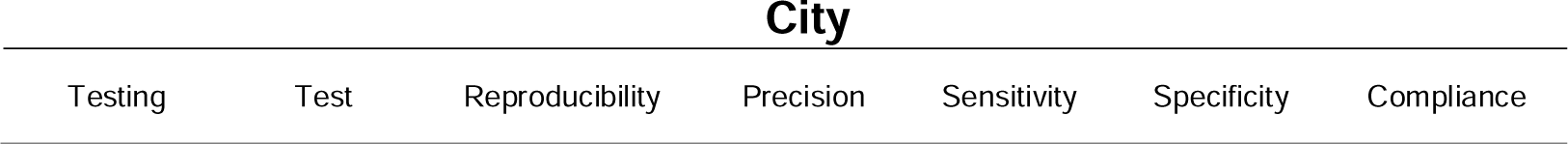
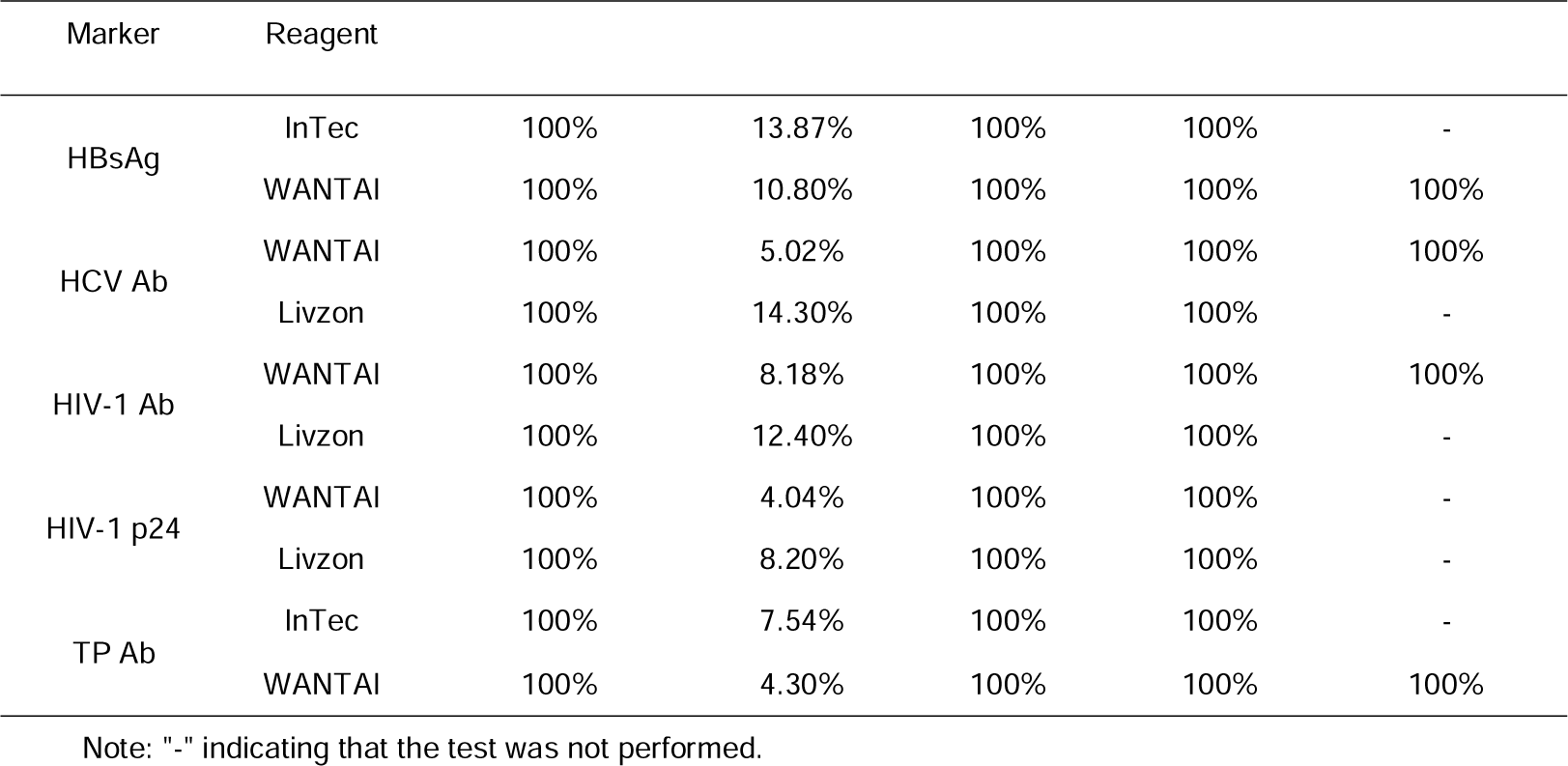
Serological Testing Performance Verification for TTIs in Nagqu City.

| Testing | Test | Reproducibility | Precision | Sensitivity | Specificity | Compliance |
| --- | --- | --- | --- | --- | --- | --- |

| Marker | Reagent |  |  |  |  |  |
| --- | --- | --- | --- | --- | --- | --- |
| HBsAg | InTec | 100% | 13.87% | 100% | 100% | - |
|  | WANTAI | 100% | 10.80% | 100% | 100% | 100% |
| HCV Ab | WANTAI | 100% | 5.02% | 100% | 100% | 100% |
|  | Livzon | 100% | 14.30% | 100% | 100% | - |
| HIV-1 Ab | WANTAI | 100% | 8.18% | 100% | 100% | 100% |
|  | Livzon | 100% | 12.40% | 100% | 100% | - |
| HIV-1 p24 | WANTAI | 100% | 4.04% | 100% | 100% | - |
|  | Livzon | 100% | 8.20% | 100% | 100% | - |
| TP Ab | InTec | 100% | 7.54% | 100% | 100% | - |
|  | WANTAI | 100% | 4.30% | 100% | 100% | 100% |

Note: "-" indicating that the test was not performed.

**Table 3.**
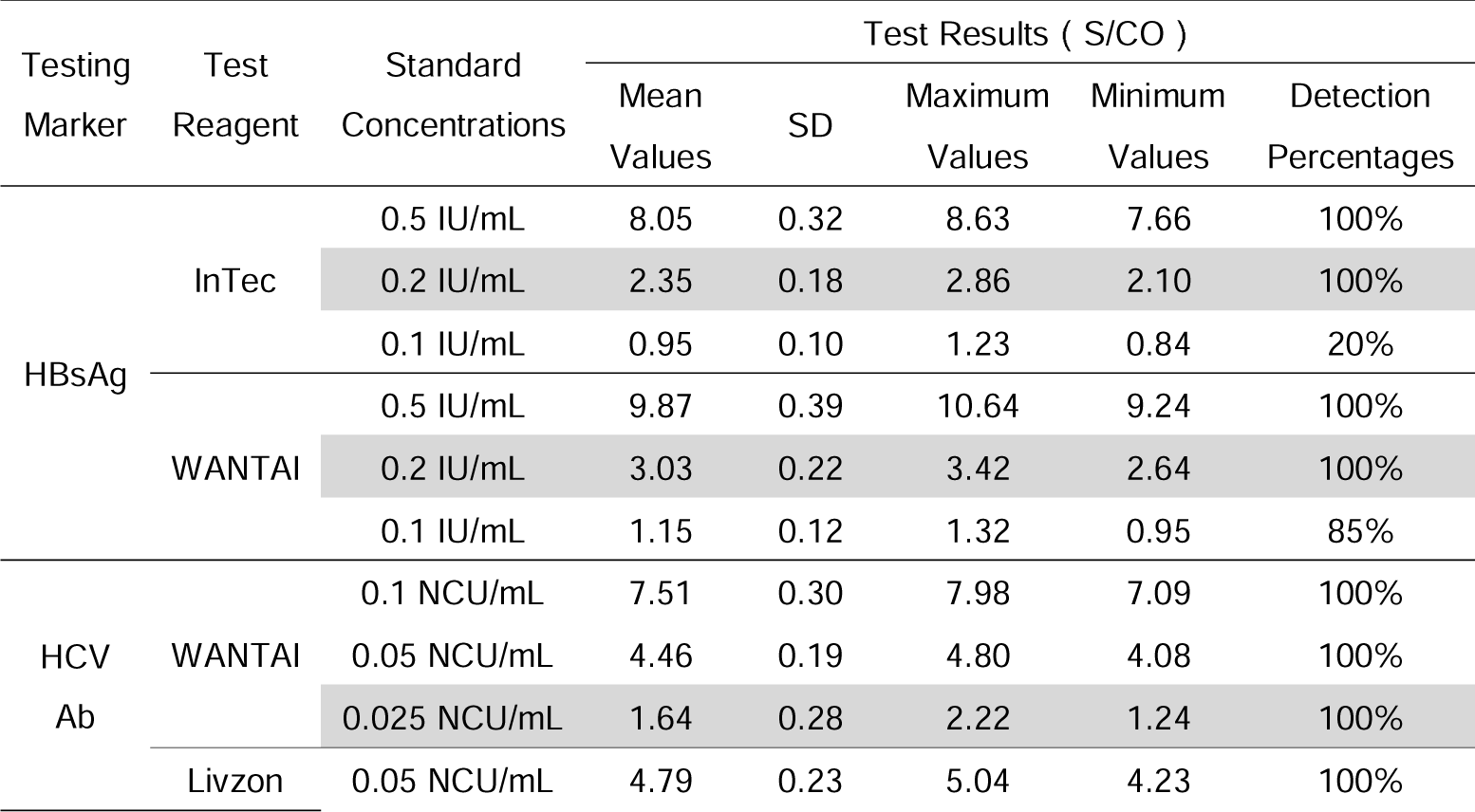
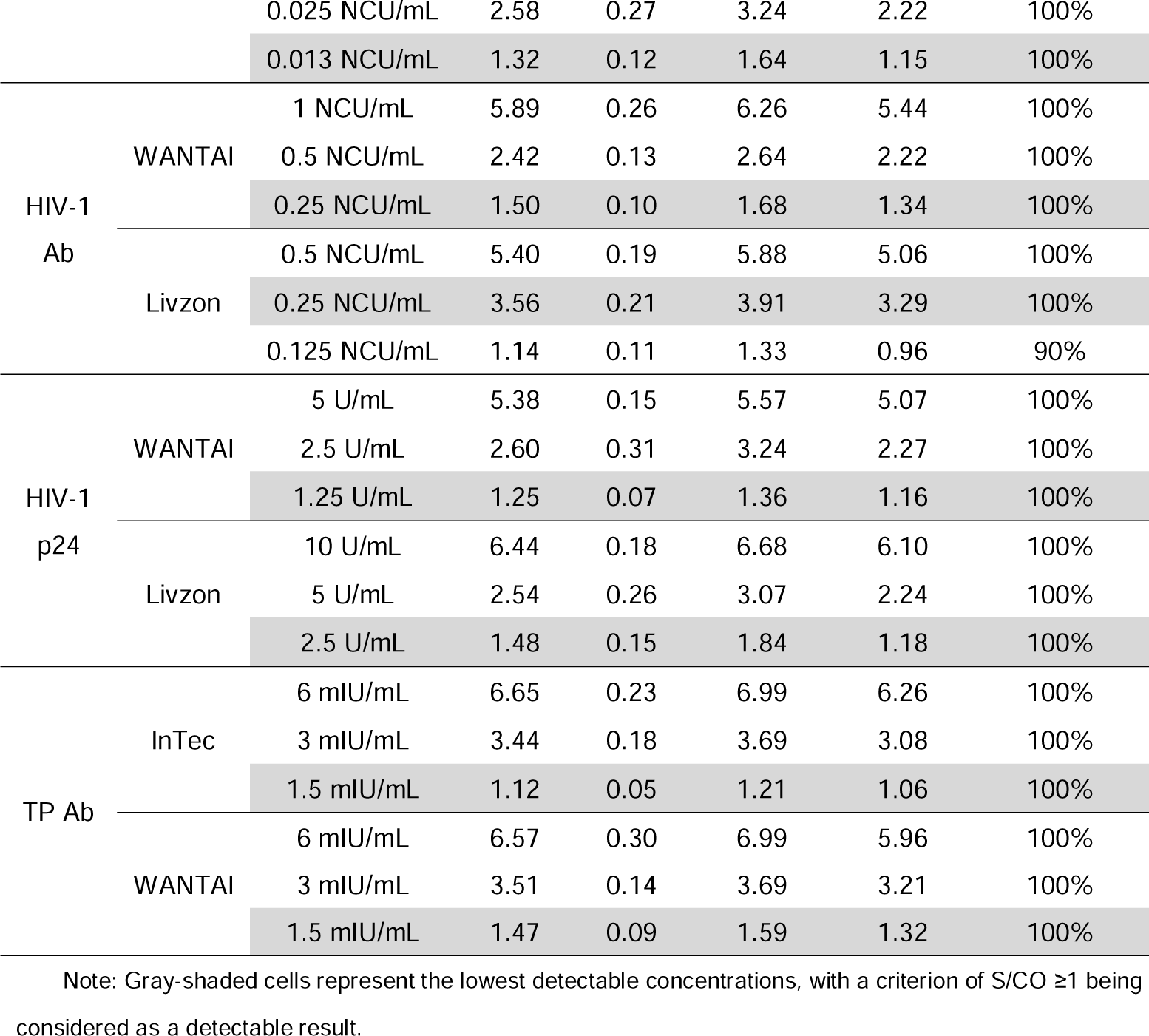
Serological Testing LOD for TTIs in Nagqu City.

**Table 4.** Performance Verification of NAT for TTIs in Nagqu City.

| TTIs<br>Markers | Genotype | Average 95% LOD<br>(95% CI) (IU/mL) | Reproducibility | Precision | Compliance |
| --- | --- | --- | --- | --- | --- |
| HBV DNA | HBV genotype C | 8.29(5.88-20.55) | 100% | 2.01% | 100% |
|  | HCV subtype 2a | 97.14(71-182.67) |  |  |  |
| HCV RNA | HCV subtype 1b | 176.64(136.41-270.98) | 100% | 3.18% | 100% |
|  | HCV subtype 6a | 198.11(149.42-320.88) |  |  |  |
| HIV-1 RNA | HIV-1 genotype CRF07_BC | 40.52(30.26-85.92) | 100% | 4.25% | 100% |

Table 2 demonstrates the Serological Testing Performance Verification for TTIs in Nagqu City. The results indicate reliable repeatability, with all markers achieving 100% consistency in repeated testing. HIV p24 Ag (WANTAI) and TP Ab (WANTAI) showed higher precision with CV values of 4.04% and 4.3%, respectively, while HCV Ab (Livzon) and HBsAg (InTec) exhibited higher CV values of 14.30% and 13.87%, but still below the acceptable limit of 15%. Sensitivity and specificity were 100% for all markers. The EQA activities in 2021 and 2022 involved multiple assessments, and all testing items, including HBsAg, HCV Ab, HIV-1 Ab, and TP Ab, achieved 100% compliance, indicating high levels of accuracy and precision in the laboratory’s testing procedures during these years.

Table 3 and Fig 2 present the LOD for serological blood screening markers. According to the results, the LOD for HBsAg detection was 0.2 IU/mL for both InTec and WANTAI. For HCV Ab detection, WANTAI and Livzon achieved LODs of 0.025 NCU/mL and 0.013 NCU/mL, respectively. The LOD for HIV-1 Ab detection with both WANTAI and Livzon was 0.25 NCU/mL. In the case of HIV-1 p24 detection, the LODs were 1.25 U/mL and 2.5 U/mL for WANTAI and Livzon, respectively. Notably, for TP Ab detection, both InTec and WANTAI exhibited relatively low LODs at 1.5 mIU/mL.

**Fig 2.** **Detection Limits of Serological Blood Screening Markers in Nagqu City**

Table 4 presents the Performance Verification of NAT for TTIs in Nagqu City. The results demonstrate reliable analytical sensitivity, with LODs within the declared limits for HBV, HCV, and HIV detection. The precision was satisfactory for most markers, with CV values below 5%, indicating good repeatability. The system exhibited excellent specificity, as cross-contamination was not observed. Additionally, the consistency analysis showed a 100% coincidence rate between the laboratory’s results and the NCCL results, confirming the accuracy and reliability of the nucleic acid testing system used in Nagqu City.

### Evaluation of Blood Screening Strategies

The evaluation of blood screening strategies aimed to assess the performance and suitability of the Nagqu NAT system for nucleic acid screening strategies. As shown in Table 5, all the samples from Serum Panel A were tested negative for HBsAg using chemiluminescent immunoassay. These samples were confirmed by the NCCL to be HBV DNA low concentration samples. Among them, 18 samples showed reactive results in the Roche NAT system, of which 8 samples were also reactive in our initial screening, resulting in a screening positive concordance rate of 44.44%. In the confirmatory retesting, 14 samples remained reactive, resulting in a confirmatory positive concordance rate of 77.78%. Similarly, in the Haoyuan NAT system, 22 samples showed reactive results, with 13 samples being reactive in our initial screening, yielding a screening positive concordance rate of 59.09%. In the confirmatory retesting, 13 samples remained reactive, resulting in a confirmatory positive concordance rate of 59.09%. Among the 38 samples that tested positive for HBcAb in serological testing, our initial screening yielded a detection rate of 47.37% (18/38), while the confirmatory retesting had a detection rate of 68.57% (24/35). Three samples were not retested due to insufficient sample volume. The results indicate the performance of the Nagqu NAT system in detecting HBV DNA and its concordance with other NAT systems.

**Table 5.** Results of s Serum Panel A of HBV DNA low viral load samples tested in different NAT systems.

| Sample ID | Testing Reagent | Test Results ( Ct ) | Nagqu NAT Results |  |  | Serological Test Results (NCCL)<br>( Chemiluminescent Assay ) |  |  |  |  |
| --- | --- | --- | --- | --- | --- | --- | --- | --- | --- | --- |
|  |  |  | Initial Test ( Ct ) | Repeat | Repeat | HBsAg | HBsAb | HBeAg | HBeAb | HBcAb |
|  |  |  |  | Test 1 ( Ct ) | Test 2 ( Ct ) |  |  |  |  |  |
| NAT A 001 | N/A | N/A | NR | NR | NR | NR | 17.16 | NR | NR | NR |
| NAT A 002 | N/A | N/A | 33.07 | 31.81 | 32.58 | NR | NR | NR | NR | NR |
| NAT A 003 | MPX | 36.8 | 36.45 | 35.62 | 36.19 | NR | NR | NR | NR | NR |
| NAT A 004 | MPX | 38.2 | NR | NR | NR | NR | 18.04 | NR | NR | R |
| NAT A 006 | MPX | 36.3 | NR | 34.55 | NR | NR | NR | NR | NR | R |
| NAT A 007 | MPX | 36.4 | 36.82 | 33.93 | 34.83 | NR | NR | NR | NR | R |
| NAT A 009 | MPX | 35.9 | 35.97 | 36.68 | NR | NR | NR | NR | R | R |
| NAT A 010 | MPX | 37.2 | NR | 35.51 | NR | NR | NR | NR | R | R |
| NAT A 011 | MPX | 35 | NR | 36.21 | 36.50 | NR | NR | NR | R | R |
| NAT A 012 | MPX | 39 | NR | NR | NR | NR | NR | NR | R | R |
| NAT A 013 | MPX | 35.9 | 34.12 | 36.11 | 34.24 | NR | 256.5 | NR | R | R |
| NAT A 014 | MPX | 30.6 | 32.02 | 31.37 | 31.79 | NR | 12.07 | NR | NR | R |
| NAT A 015 | MPX | 34.9 | 33.86 | 35.73 | 35.57 | NR | NR | NR | R | R |
| NAT A 016 | MPX | 37.2 | NR | NR | NR | NR | NR | NR | NR | R |
| NAT A 017 | MPX | 30.4 | 33.14 | 32.05 | 31.53 | NR | NR | NR | R | R |
| NAT A 018 | MPX | 36.5 | NR | 37.14 | NR | NR | NR | NR | NR | R |
| NAT A 019 | MPX | 34.2 | 38.65 | 38.82 | 37.36 | NR | NR | NR | R | R |
| NAT A 034 | MPX | 36.3 | NR | 36.77 | 36.45 | NR | NR | NR | R | R |
| NAT A 037 | MPX | 37.8 | NR | NR | 36.33 | NR | NR | NR | R | R |
| NAT A 044 | MPX | 34.9 | NR | N/A | N/A | NR | NR | NR | R | R |
| NAT A 005 | Haoyuan | 37.8 | NR | NR | 36.16 | NR | NR | NR | NR | NR |
| NAT A 008 | Haoyuan | 35.61 | 37.67 | 34.01 | 36.83 | NR | NR | NR | R | R |
| NAT A 020 | Haoyuan | 41.53 | NR | NR | NR | NR | NR | NR | R | R |
| NAT A 021 | Haoyuan | 41.37 | NR | NR | NR | NR | NR | NR | NR | R |
| NAT A 022 | Haoyuan | 44.06 | NR | NR | NR | NR | 16.68 | NR | R | R |
| NAT A 023 | Haoyuan | 40.74 | 36.37 | 36.68 | 35.33 | NR | NR | NR | NR | R |
| NAT A 024 | Haoyuan | 40.91 | 36.14 | NR | NR | NR | NR | NR | R | R |
| NAT A 025 | Haoyuan | 41.3 | 37.11 | 35.85 | 35.74 | NR | NR | NR | NR | R |
| NAT A 026 | Haoyuan | 38.31 | NR | 44.19 | 36.72 | NR | NR | NR | R | R |
| NAT A 027 | Haoyuan | 43.982 | NR | NR | NR | NR | NR | NR | NR | R |
| NAT A 028 | Haoyuan | 43.557 | NR | NR | NR | NR | 88.99 | NR | NR | R |
| NAT A 029 | Haoyuan | 40.138 | 36.62 | 39.43 | 35.90 | NR | NR | NR | NR | R |
| NAT A 030 | Haoyuan | 38.304 | 37.08 | 35.42 | 35.98 | NR | NR | NR | R | R |
| NAT A 031 | Haoyuan | 42.05 | NR | NR | 37.36 | NR | 68.66 | NR | R | R |
| NAT A 032 | Haoyuan | 33.3 | 33.23 | 32.20 | 32.34 | NR | 37.8 | NR | NR | R |
| NAT A 033 | Haoyuan | 41.35 | 44.29 | 36.14 | NR | NR | NR | NR | NR | NR |
| NAT A 035 | Haoyuan | 39 | 37.50 | NR | 35.91 | NR | NR | NR | R | R |
| NAT A 036 | Haoyuan | 36.22 | 35.03 | 34.42 | 34.59 | NR | NR | NR | R | R |
| NAT A 038 | Haoyuan | 38.57 | NR | 36.10 | 36.40 | NR | NR | NR | NR | R |
| NAT A 043 | Haoyuan | 38.65 | 37.95 | NR | NR | NR | NR | NR | NR | NR |
| NAT A 045 | Haoyuan | 37.45 | 35.90 | N/A | N/A | NR | NR | NR | NR | R |
| NAT A 046 | Haoyuan | 42.81 | 33.28 | N/A | N/A | NR | 87.2 | NR | R | R |

Note: The sample IDs used in this study were strictly internal and confidential, known only within the research group.
MPX: cobas® TaqScreen MPX Test Kit (v2.0) tested by Roche cobas s 201system. Haoyuan: Haoyuan Biotech NAT
Kit for HBV/HCV/HIV -1 ((Real-time PCR) tested by Haoyuan ChiTaS BSS1200 NAT system. R: Reactive. NR:
Non-reactive. N/A: Untested. Parallel NAT system detection results data provided by NCCL.

The results demonstrate the effectiveness of the nucleic acid testing system, particularly in detecting low viral load HBV infections. The system’s adoption has proven to be more efficient in preventing the transmission of HBV through blood transfusions in high-prevalence regions like Nagqu City. The findings underscore the importance of NAT in maintaining a safe blood supply and reducing the risk of transfusion-transmitted HBV infections. Moreover, the testing strategy is also cost-effective and suitable for high-altitude regions with low donation volume, using a single-test mode. Despite the challenges posed by the high-altitude and dry climate of Nagqu City, the blood screening procedures have demonstrated minimal differences compared to the plains regions. The system exhibited excellent stability and accuracy, ensuring reliable blood screening results even under such environmental conditions.

## Discussion

This study aimed to investigate the blood screening strategies in Nagqu City, Tibet, with a specific emphasis on the performance of serological and nucleic acid testing. Despite encountering distinctive challenges of geographical remoteness, high-altitude hypoxia, and a dry climate, valuable insights can be gleaned from the analysis of blood screening outcomes.

The effectiveness and reliability of blood screening have demonstrated continuous improvement, as evidenced by a declining trend in the rate of unqualified donations over time, indicating the enhanced effectiveness of screening strategies (Table 1). However, it is crucial to note that Nagqu City’s rates of unqualified donations for various markers still exceed the national average. The majority (58.13%, 143/246) of the unqualified blood donations detected in laboratories were attributed to failing the ALT screening, which may be related to the donors’ liver function. However, it is more likely associated with unhealthy lifestyle habits, such as poor diet, alcohol consumption, and medication use. To ensure the effective utilization of blood resources, the importance of pre-donation screening for ALT should be emphasized. Notably, positive results for HBV were the primary reason for donor deferral in TTIs tests, with 9.76% (24/246) attributed to HBsAg and 11.38% (28/246) to HBV-DNA. Additionally, Treponema pallidum (TP), which causes syphilis, accounted for a significant proportion (15.85%, 39/246) of unqualified donations. These findings align with prevailing trends observed in other high-altitude regions of the Qinghai-Tibet Plateau but are considered considerably high when compared to other developing countries[15–18]. Moreover, it is essential to acknowledge that the prevalence of HBV infection remains alarmingly high in China, with approximately 100 million people suffering from chronic HBV infection[19]. The high prevalence of HBV and other infectious markers in the blood donor pool in this region emphasizes the importance of stringent quality management and individual donation nucleic acid detection protocols in blood banks to ensure blood safety and prevent potential HBV transfusion transmission[20–22]. Efforts should be directed towards implementing effective measures to reduce the number of unqualified donations, continuing to monitor the local TTIs’ prevalence trends, and enhancing overall blood safety in this geographically challenging area[23, 24]. By addressing these challenges and implementing targeted interventions, the blood screening system in Nagqu City can further improve its effectiveness and contribute to safer and more reliable blood donations.

The challenging high-altitude, oxygen-deficient, and dry climate conditions in Nagqu indeed present unique obstacles to the blood screening process. However, the results of the performance verification (as detailed in Tables 2, 3 and 4) illustrate the reliability and accuracy of both serological and nucleic acid testing procedures in overcoming these challenges. Across all markers, there is a consistent demonstration of high levels of consistency, reproducibility, sensitivity, and specificity, ensuring that results are both reliable and precise. One of the critical parameters in evaluating the effectiveness and sensitivity of diagnostic assays is the LOD for serological blood screening markers. This study undertook a comprehensive investigation into the LODs for various bloodborne pathogens, encompassing HBsAg, HCV Ab, HIV-1 Ab, HIV-1 p24, and TP Ab, using different test reagents. Remarkably, the observed LOD of 0.2 IU/mL for HBsAg detection in Nagqu City stands out, as it is significantly lower than the LODs reported in the literature for domestically produced test reagents. Typically, these reported LODs are around 0.6-1 IU/mL [25, 26]. This remarkable increase in sensitivity is particularly advantageous, especially in regions where early detection of hepatitis B infection is of paramount importance due to the high prevalence of this disease. The findings shed light on the performance and capabilities of these assays in Nagqu City, Tibet, under challenging environmental conditions.

In this study, the NAT system displayed excellent capability in detecting HBV DNA, which is particularly beneficial in high-altitude regions with a high prevalence of HBV transmission[27]. The adoption of the NAT system effectively detects low viral load HBV infections, minimizing the risk of HBV transmission through blood transfusions[28]. This finding is crucial in reducing the risk of transfusion-transmitted HBV infections and emphasizes the significance of NAT in maintaining a safe blood supply. Other regions can learn from Nagqu City’s NAT strategies to strengthen monitoring and prevention measures for low viral load infections, further enhancing transfusion safety.

The research findings indicate minimal differences in the blood screening process between Nagqu and plains regions in China[29–33]. Furthermore, the system demonstrated exceptional stability and accuracy, signifying significant progress in adapting to the high-altitude and dry conditions. The adaptability and performance of the blood screening system in challenging environmental conditions are commendable, highlighting the commitment of healthcare professionals in Nagqu City to ensuring safe and reliable blood donations. Despite the unique challenges posed by the high-altitude and dry climate, the blood screening process in Nagqu City has demonstrated remarkable effectiveness and reliability. The successful implementation of the NAT system and the consistent performance of serological testing underscore the commitment to maintaining a safe blood supply and preventing transfusion-transmitted infections. These research findings not only benefit Nagqu City but also provide valuable insights and practical guidance for other high-altitude regions facing similar challenges in blood screening and transfusion safety.

In conclusion, this study provides a comprehensive evaluation of blood screening strategies in Nagqu City, demonstrating the effectiveness and reliability of the current procedures. The research also offers valuable insights and practical implications for other geographically remote and high-altitude regions, particularly in nucleic acid testing. Through continuous improvement and optimization strategies, we can further enhance the quality of blood screening, ensure transfusion safety, and provide beneficial experiences and guidance for blood screening in similar regions. This study fills gaps in knowledge and offers important references and practical value for improving blood screening and enhancing transfusion safety in geographically remote regions. Future research should explore blood screening strategies adapted to high-altitude and dry conditions to provide broader significance for other geographically remote regions.

## Data Availability

All relevant data are within the manuscript and its Supporting Information files.

## Acknowledgments

We extend our heartfelt gratitude to the National Center for Clinical Laboratories their invaluable support, including their high regard for this study, provision of panel samples, and technical assistance. Our sincere appreciation goes to the Tibetan government for their multifaceted support. Special recognition is due to the dedicated staff members of the blood bank laboratories, whose unwavering support during the experiments is highly commendable.

## Author Contributions

Conceptualization: Yaxin Fan

Data curation and analysis: Liang Zang, Lei Zhou, Ji Duo

Investigation: Liang Zang, Lei Zhou, Yaxin Fan, Xiaohua Liang, Ji Duo

Methodology: Liang Zang, Lei Zhou, Yaxin Fan, Xiaohua Liang, Ji Duo, Hao Lin, Rui Bai, Mei Yang, Chao Dan

Project Administration: Liang Zang, Yaxin Fan, Xiaohua Liang

Resources: Liang Zang, Yaxin Fan

Supervision: Yaxin Fan, Xiaohua Liang

Writing: Liang Zang

Writing - Review & Editing: Liang Zang, Lei Zhou, Yaxin Fan, Xiaohua Liang, Ji Duo, Hao Lin, Rui Bai, Mei Yang, Chao Dan.

